# Type and frequency of ocular and other known symptoms experienced by people who self–diagnosed as suffering from COVID-19 in the UK

**DOI:** 10.1101/2020.06.20.20134817

**Authors:** Shahina Pardhan, Megan Vaughan, Jufen Zhang, Lee Smith, Havovi Chichger

## Abstract

**Background:** Recent literature suggests that ocular manifestations present in people suffering from COVID-19. However, the prevalence and the type of ocular symptoms varies substantially, and most studies report retrospective data from patients suffering from more serious versions of the disease. Little is known of exactly which ocular symptoms manifest in people with milder forms of COVID-19.

**Methods:** An online questionnaire obtained self-report data from people in the community, who reported to be inflicted with COVID-19. The type and frequency of different symptoms suffered during COVID-19 were obtained. Details of any pre-existing ocular conditions and the duration of symptoms of COVID-19 were ascertained.

**Results:** Data from 132 participants showed that the four most reported COVID-19 symptoms were Dry Cough (63%), Fever (67%), Fatigue (83%), and loss of Smell/Taste (63%). 56% of the participants reported to having experienced an eye symptom, 46% reported to having a new or different eye symptom compared to pre-COVID-19 state. Three ocular symptoms (watery eyes, sore eyes, sensitivity to light) were significantly different from Pre-COVID-19 state (p<0.05). Logistic regression showed a significant association of eye symptoms with Fever (p=0.035).

**Conclusion:** Nearly half of the sample of people studied experienced ocular symptoms. The significant ocular symptoms, indicative of viral conjunctivitis, might have been missed in patients with more serious manifestations of the disease. It is also important to differentiate between the types of ocular manifestation, as symptoms of bacterial conjunctivitis (i.e. mucous discharge, gritty eyes) were not significant. Possible mechanisms for SARS-CoV-2 infection within the eye are discussed.

**Key message:** To date, there are no studies on ocular symptoms experienced by people who self-reported as suffering from mild case of COVID-19. In the absence of population –wide testing in the UK, our study shows that nearly half of the population reported to having experienced an eye symptom. It is likely that the significant ocular symptoms, indicative of viral conjunctivitis, might have been overlooked in the light of other more serious and critical manifestations of COVID-19. The data are important, especially in countries that rely on self –report of COVID-19 symptoms when confirmative tests for COVID-19 are not available.

## INTRODUCTION

SARS-CoV-2 is the cause of COVID-19 which has been shown to be primarily a respiratory illness with the most common symptoms being a new, continuous cough, and high temperature. Further symptoms have been reported, with the World Health Organisation (WHO) adding conjunctivitis to the less common symptoms.[1] Ocular manifestations of COVID-19 have not been at the forefront of the substantial research, possibly due to the life-threatening nature of the other more serious respiratory symptoms which have resulted in high numbers of mortality around the world. This might have resulted in other non-life-threatening symptoms not being given much importance when patients present with life threatening conditions. However, the types, frequency and subsequently, the transmissibility of the virus via ocular secretions must be considered, especially since the eye, with the nose and mouth, has been recognised as one of the organs through which the virus might enter the body.

In the absence of wide-spread testing for infection, UK’s government advice was that anybody who experienced the known symptoms of COVID-19, such as: dry, continuous cough and fever, should self-isolate. This was also extended to family members and other people living in the same house. Testing was reserved for those experiencing significant symptoms. As such, a true documentation of signs and symptoms of those with the virus is limited. Self-reported ocular symptoms appeared prior to respiratory signs, in a number of cases.[2, 3]

The reports on ocular manifestation is varied and shows a wide range in terms of prevalence. A recent review by Ulhaq and Soraya, reported a low prevalence of ocular symptoms amongst those with a confirmed case of the virus.[4] These range from 5% to 31% in confirmed cases and it is possible that ocular manifestations were not considered in the presence of other more critical symptoms when patients were admitted to hospital. A study that examined 56 patients showed that 27% (n=15) of confirmed COVID-19 patients reported ocular symptoms: sore eyes, itching, foreign body sensation, hyperaemia, floaters and/or secretions (although the type of secretions was not noted), with six patients reporting ocular symptoms prior to respiratory or fever symptoms onset.[5] A larger study in China on 534 confirmed cases reported only 25 cases (5%) with ocular manifestations.[6] They reported three main ocular symptoms as dry eye, foreign body sensation and blurred vision, and similar to the aforementioned study a small proportion of patients (n=3) reported ocular symptom(s) as their first symptom. In both studies, reported a positive association was shown between the disease severity and the prevalence of ocular manifestations.[5, 6] Recently, late onset (>2 weeks post initial influenzae symptoms) ocular manifestations were reported in a confirmed COVID-19 patient (in France) who displayed pseudomembranous and haemorrhagic conjunctivitis.[7] However conjunctival swabs were returned as negative for both a bacterial and viral manifestation. Other studies have reported the presence of SARS-CoV-2 virus in conjunctival swabs in people suffering of ocular manifestations, and also in those who did not actually manifest with conjunctivitis symptoms.[2, 8]

Initially, ocular transmission was not thought to be a possibility. However a study from a previous coronavirus outbreak (MERS-CoV) showed that those who wore full protective wear, including eye protection, showed no evidence of serum antibodies.[9] Reports of full personal protective equipment without eye protection, has been linked to the spread of the SARS-CoV-2 with reports of unilateral conjunctivitis, and a fever following later, which appears to mirror the new SARS-CoV-2.[8, 10]

It is also possible that the virus is present in the ocular surface but in the absence of any ocular signs, no conjunctival swabs were considered necessary to be taken.[8] In addition, the majority of ocular manifestations of the virus are self-limiting, and as they are not as serious as the other life threatening symptoms of COVID-19, it is possible that they may be under-reported in the light of other more serious symptoms.[11]

To date very little is known about the manifestation of the type of ocular symptoms (watery eyes, sore eyes etc) that are most experienced by people in the UK suffering from COVID-19 in people in the UK who were not hospitalised. There is also little known on how these ocular manifestations relate to the other known COVID-19 like symptoms (such as fever and dry cough). The aims of the study were to therefore determine the type and frequency of ocular symptoms, compared to other symptoms through self-report by people experiencing COVID-19 symptoms in the UK.

## MATERIALS AND METHODS

### Methods

An online questionnaire was developed to examine the type and frequency of ocular and other symptoms in people who had suffered from COVID-19. The questionnaire ascertained the type and frequency of different already known symptoms of COVID-19 (dry cough, fever, fatigue, loss of smell/taste), as well as of different ocular symptoms (sore eyes, watery eyes, pain in the eye). The time window of how long these symptoms were experienced were also compared with other known symptoms of COVID-19. Data on consultation with healthcare practitioners and self-isolation were also determined.

The questionnaire also determined if participants were chronic sufferers of the ocular symptoms (pre COVID-19) and if they had experienced the same ocular symptoms within three weeks of the other COVID-19 symptoms. The questionnaire disseminated through word of mouth, leaflets, websites and other avenues including via social media, and invited people who self -reported having suffered from any COVID-19 symptoms to take part.

### Participants

Data were collected between 16th April 2020 and 15th May 2020. Eligible data from one hundred thirty-two participants (n=132) over the age of 18 years and living in the UK who self -reported having suffered from COVID-19 symptoms were analysed. All patients provided informed consent for taking part in the study. Ethical approval was granted by the Faculty of Health, Education, Medicine and Social Care Ethics Committee at Anglia Ruskin University. All patients provided informed consent prior to completing the questionnaire. Participants were treated in accordance with the Declaration of Helsinki.

It was not appropriate or possible to involve patients or the public in the design, or conduct, or reporting, or dissemination plans of our research

## Results

Table 1 shows the questions and summary data of the participants (n=132) who completed the survey.

**Table 1:** Date of survey: 16th April 2020 to 15th May 2020.

|  |
| --- |
| Participants (n=132) could only access the survey if they self-reported they were 18 years |
| or older and had experienced symptoms indicative of COVID-19 |
| 1. What is your Age Group?<br>Summary data: 18-29 years (14%), 30-39 years (25%), 40-49 years (32%), 50-59 years (17%), 60-69 years (9%), 70-79 years (3%), over 80 years (0%) |
| 2. What is your Gender?<br>Summary data: Males (37%), Females (63%) |
| 3. What Symptoms indicative of COVID-19 did you experience?<br>Summary data:<br>Fatigue (83%), Fever (68%), Dry Cough (63%), Loss of Smell /Taste (63%). Others included Sore throat (53%), Diarrhoea (22%), Headaches (17%), Shortness of Breath (17%), Myalgia/other pain (10%). |
| 4. Do you suffer from any systemic diseases?<br>Summary data: Hypertension, diabetes, Asthma, Lung disease (27%), No systemic disease (73%) |
| 5. Did you experience any eye symptoms when you experienced other symptoms indicative of COVID-19?<br>Summary data 56% of participants reported to having experienced an eye symptom during COVID-19. 46% of those who reported 'yes' had reported a new or a different eye symptom, see Table 2. |
| 6. How long did you experience the symptoms indicative of COVID-19?<br>Summary data: COVID-19 symptoms: less than one week (30%), Between 1-2 weeks (40%), over two weeks (30%) |
| 7. How long did you experience the symptoms indicative of COVID-19?<br>Summary data: Eye Symptoms: less than one week (43%), Between 1-2 weeks (42%), over two weeks (15%), see Table 2. |
| 8. When did you experience the eye symptoms (compared to other symptoms indicative of COVID-19)?<br>Summary data: 3 weeks before (7%), within 1-2 weeks (19%), (within a week (59%), one week after (15%) |
| 9. Who did you consult about your eye problems during COVID-19?<br>Summary data: Doctor (24%), Optometrist (16%), Pharmacist (10%), |
| 10. If you self-isolated- how long did you isolate for? |
| Summary data: 1 week (21%), 2 weeks (40%), longer 2 weeks (29%), did not isolate/isolated/did not respond (10%). |

### Statistical analysis

The association between two categorical variables (i.e. gender) was assessed using the Chi-square test. McNemar’s test compared the difference between paired nominal data for all the symptoms and also for individual eye symptoms pre-COVID-19 and during COVID-19. Multivariate logistic regression models reporting odds ratio (OR) and the 95% confidence intervals were used to evaluate the association between the systemic condition (dry cough, fever, loss of smell/taste, fatigue), and eye symptoms during COVID-19. Statistical tests were performed at the two-sided significance level of 0.05. The Stata statistical package version 14.2 was used to analyse the data.

### COVID-19 symptoms

95% of the participants experienced at least one of the four most common COVID-19 symptoms experienced (Fatigue, Fever, Dry Cough, Loss of Smell /Taste) (Table 1). It should be noted that more than one symptom was experienced by the majority of participants.

### Ocular symptoms

56% of the participants reported to having experienced an eye symptom, 46% reported to having a new or different eye symptom compared to pre-COVID-19 state. The most frequent ocular symptoms included watery eyes (16%), sore eyes (19%), itchy eyes (15%), photophobia (18%). Table 2 compares ocular symptoms pre and during COVID-19 using McNemar’s test for paired comparisons. Three ocular symptoms (watery eyes, sore eyes, photophobia) were shown to be significantly different (p<0.05).

**Table 2:** A comparison between pre COVID19 and during COVID-19.

|  | Pre<br>COVID-19 | During COVID-19 | McNemar's<br>Paired test. |
| --- | --- | --- | --- |

|  | (n=132) | (n=132) | p-value |
| --- | --- | --- | --- |
| All eye symptoms | 55 (41.7) | 74 (56.1) | 0.012 |
| Dry eye (ns) | 27 (20.5) | 18 (13.6) | 0.06 |
| Watery eyes (s) | 9 (6.8) | 21 (15.9) | 0.014 * |
| Mucous discharge (ns) | 2 (1.5) | 8 (6.1) | 0.06 |
| Changes in the eyelids (ns) | 4 (3.0) | 1 (0.8) | 0.18 |
| Gritty eyes (ns) | 4 (3.0) | 9 (6.8) | 0.13 |
| Sore eyes (s) | 4 (3.0) | 25 (18.9) | <0.001 * |
| Itchy eyes (ns) | 18 (13.6) | 20 (15.2) | 0.68 |
| Foreign body sensation (ns) | 4 (3.0) | 5 (3.8) | 0.70 |
| Photophobia | 13 (9.8) | 25(18.9) | 0.0186 * |

### Duration of time frame for COVID-19 and eye symptoms

Chi –square showed no significant differences in the duration of eye symptoms and the other symptoms of COVID-19 (p=0.164).

### Differences between males and females

Chi square analysis showed no significant in ocular symptoms between males and females (p=0.235)

### Association of Eye symptoms and the other COVID-19 symptoms

Table 3 shows multivariable logistic regression model’s outputs, evaluating the association between the eye symptoms and the other COVID-19 symptoms. Fever was significantly associated with eye symptoms (OR 2.29; 95% CI 1.06 to 4.94)

**Table 3:** Multivariable logistic regression models of risk factors for eye symptoms (n=132)

| Variables | OR (95% CI) | p-value |
| --- | --- | --- |
| Dry cough | 0.61 (0.28-1.33) | 0.21 |
| Fever | 2.29 (1.06-4.95) | 0.034 |
| Smell/test | 1.09 (0.51-2.38) | 0.82 |
| Fatigue | 1.42 (0.54-3.75) | 0.48 |
| Constant | 0.71 (0.27-1.88) | 0.49 |

## DISCUSSION

In the present sample of the UK public who were inflicted with COVID-19 symptoms, approximately half of the sample reported experiencing new or different eye symptoms. The particular type of ocular symptoms that were shown to be significant are indicative of viral conjunctivitis. Although ‘conjunctivitis’ is included in the list of less common symptoms by the WHO, it is important to differentiate between symptoms shown here of other eye conditions, such as bacterial conjunctivitis which manifests mainly as mucous discharge or gritty eyes. It is also likely that the ocular symptoms of viral conjunctivitis might have been overlooked in the light of other more serious and critical manifestations of COVID-19.

The prevalence of the four most reported COVID-19 symptoms in our study, dry cough (63%), fever (67%), fatigue (83%), and loss of smell/taste (63%), agree with data from prevalence studies carried out on larger samples. Also, the ocular symptoms were experienced within the same timeframe as other COVID-19 symptoms, indicating that the eye was affected around the same time as other parts of the body.

The potential mechanism for SARS-CoV-2 infection in the eye is important. SARS-CoV-2 invasion of healthy human cells is reliant on the host receptor, ACE2, hypothesised to infect cells using two potential routes.[12] The traditional route of entry is through the spike protein (S) of the virus which binds to ACE2 receptor as a homodimer.[13] The S-protein is then cleaved by the transmembrane protease, TMPRSS2, into S1 and S2 subunits.[14] The latter is responsible for membrane fusion, to allow entry into the cell via cathepsin L and cathepsin B mediated endocytosis.[15, 16] An alternate hypothesised route for SARS-CoV-2 infection into human cells is the ability to bind to the ACE2-B°AT1 heterodimeric complex at the human cell surface).[13] B°AT1 (SLC6A19), traditionally considered to be an amino acid transporter in the small intestine, has gained significant interest as ACE2 is also responsible for the membrane trafficking of B°AT1.[17] Whilst ACE2 and TMPRSS2 expression has been studied in the eye, given the early stage of studies on B°AT1 in relation to SARS-CoV-2 infection, the protein has not yet been identified in the eye.

SARS-CoV-2 is typically considered to be transmitted by air-borne dissemination of respiratory droplets through direct or indirect contact. Many viruses such as avian influenza virus H7 have been shown to cause highly infectious viral conjunctivitis, and conjunctiva is hypothesised to be an important entry point for the infection.[18] Although the data presented here are the first data to report these findings outside hospital settings in the UK, previous studies in China demonstrate that up to a third of patients with COVID-19 have suffered from ocular conditions associated with conjunctivitis, such as watery, and sore eyes.[5, 6, 19] Whilst there are no studies, as yet, that have determined conclusively the mechanism through which SARS-CoV-2 can infect the conjunctiva, the eye is known to have an internal (aqueous humuor, iris, retina), and external (conjunctiva, cornea) intra-ocular renin-angiotensin system.[20] There is still controversy in the literature regarding the presence of the machinery needed for SARS-CoV-2 infection in the conjunctiva. Some studies have reported that an expression of ACE2 and TMPRSS2 in the human conjunctival and pterygium cell lines, and tissue.[21, 22] Whilst others show negligible ACE2 expression in the human conjunctiva.[23] There is therefore a great need to further investigate the possibility that SARS-CoV-2 can directly infect the conjunctiva and cause the ocular symptoms we observe in participants in this study. Another possibility is that the cornea is the site of SARS-CoV-2 infection. In cornea limbal stem cells from healthy human participants and murine cornea, high mRNA expression of ACE2 and TMPRRS2 have been identified, suggesting that SARS-CoV-2 may infect the ocular surface via the cornea, using the traditional ACE2-TMPRSS2-mediated mechanism of cell entry.[21, 22, 24] It is therefore possible that SARS-CoV-2 can infect the ocular surface via the cornea, using the traditional ACE2-TMPRSS2-mediated mechanism of cell entry. Given the neuronal expression of ACE2 and TMPRSS2, it is possible that this type of infection may allow the spread of the virus through the nose, lungs, blood stream, and through the nervous system (via the trigeminal nerve) to potentially cause the COVID-19 symptoms documented in participants.[25]

There is a strong association between the neuroinvasive potential of SARS-CoV-2 and the onset of respiratory failure in patients with COVID-19.[26] In both symptomatic and asymptomatic patients with SARS-CoV-2, nasal swabs have a significantly higher viral load than throat swabs. In addition, similar viruses to SARS-CoV-2 have been found in tears of patients infected with the virus.[27] As dry cough is one of the predominant observed symptoms in participants, and that ocular manifestations occur simultaneously with other COVID-19 symptoms, another possibility is that lacrimal drainage from the conjunctival sac into the nasal cavity allows the spread of SARS-CoV-2 into the upper respiratory tract as a potential mechanism of virus spread. In addition, in keeping with other studies, we demonstrate a large number of participants with COVID-19 symptoms indicate a loss of smell and taste.[28-30] This is not surprising given the association between viral infection and/or upper respiratory tract infections and ageusia, and anosmia.[31] Interestingly, the machinery for the main route of entry for SARS-CoV-2, ACE2 and TMPRSS2, have been identified to colocalise with the epithelium in the oral and nasal cavity where taste and smell are governed respectively.[32, 33] It is therefore possible that the spread of SARS-CoV-2 through lacrimal drainage of the tears, enables the virus to bind to ACE2 in the oral and nasal cavity to blunt taste and smell.

This is the first study to investigate eye symptoms in relation to COVID-19 in a sample of the UK population. However, findings must be interpreted in light of the study limitations. Although the number of people in this study sample is relatively small and may not be representative of the population in the UK, the prevalence of other symptoms of COVID-19 agree with those in the literature. It is also possible that some of the responders may have had other diseases /infections and not COVID-19. However, in the absence of community-based swab tests or antibody tests which were not readily available to the population of UK, we employed a criterion of including only those responders who had experienced at least two of the well-known COVID-19 symptoms. In addition, a substantial percentage of the responders (90%) had self-isolated due to the possibility of having suffered from COVID-19. It is possible that some of the ocular symptoms may have been due to other eye infections and not COVID-19 but the ocular symptoms were experienced within the same time frame as the other COVID-19 symptoms, strongly suggesting a connection to the disease.

In conclusion, findings from this UK sample found that the significant ocular manifestations of COVID-19 (sore eyes, watery eyes, photophobia) are similar to those experienced with viral conjunctivitis. This should be confirmed with a larger scale study. It is also important to note that ocular symptoms might have been under-reported in the presence of other, more serious manifestations of the disease. These findings are important for countries that also rely on self–reporting of COVID-19 symptoms when confirmative tests for COVID-19 are not available.

## Data Availability

Page 6 in attached text

## DECLARATIONS

### Contributors

SP and HC conceived and designed the study. SP and JF analysed the data. SP, MV, HC and LS wrote and revised the paper.

